# Early start of oral clarithromycin is associated with better outcome in COVID-19 of moderate severity: the ACHIEVE open-label trial

**DOI:** 10.1101/2020.12.22.20248753

**Authors:** Konstantinos Tsiakos, Antonios Tsakiris, Georgios Tsibris, Pantazis Voutsinas, Periklis Panagopoulos, Maria Kosmidou, Vasileios Petrakis, Areti Gravvani, Theologia Gkavogianni, Eleftherios Klouras, Konstantina Katrini, Panagiotis Koufargyris, Iro Rapti, Athanassios Karageorgos, Emmanouil Vrentzos, Christina Damoulari, Vagia Zarkada, Chrysanthi Sidiropoulou, Sofia Artemi, Anastasios Ioannidis, Androniki Papapostolou, Evangelos Michelakis, Maria Georgiopoulou, Dimitra-Melia Myrodia, Panteleimon Tsiamalos, Konstantinos Syrigos, George Chrysos, Thomas Nitsotolis, Haralampos Milionis, Garyphallia Poulakou, Evangelos J. Giamarellos-Bourboulis

## Abstract

**Background:** To study the efficacy of oral clarithromycin in moderate COVID-19.

**Methods:** An open-label non-randomized trial in 90 patients with COVID-19 of moderate severity was conducted between May and October 2020. The primary endpoint was defined at the end-of-treatment (EOT) as no need for hospital re-admission and no progression into lower respiratory tract infection (LRTI) for patients with upper respiratory tract infection; and as at least 50% decrease of the respiratory symptoms score the without progression into severe respiratory failure (SRF) for patients with LRTI. Viral load, biomarkers, the function of mononuclear cells, and safety were assessed.

**Results:** The primary endpoint was attained in 86.7% of patients treated with clarithromycin (95% CIs 78.1-92.2%); this was 91.7% and 81.4% among patients starting clarithromycin the first 5 days from symptoms onset or later (odds ratio after multivariate analysis 6.62; p: 0.030). The responses were better for patients infected by non-B1.1 variants. Clarithromycin use was associated with decreases in circulating C-reactive protein, tumour necrosis factor-alpha and interleukin (IL)-6; by increase of Th1 to Th2 mononuclear responses; and by suppression of SARS-CoV-2 viral load. No safety concerns were reported.

**Conclusions:** Early clarithromycin treatment provides most of clinical improvement in moderate COVID-19 (**Trial Registration:** ClinicalTrials.gov, NCT04398004)

## INTRODUCTION

Early at the beginning of the COVID-19 pandemic azithromycin, a macrolide drug was introduced in the treatment algorithm due to its anti-inflammatory properties [1, 2]. The anti-inflammatory properties of macrolides for the empirical management of community-acquired pneumonia (CAP) are reflected in the guidelines of the American Thoracic Society published in 2019 [3]. These guidelines are influenced by observational studies showing that the addition of a macrolide decreases the risk of death from severe CAP [4-8]. Two double-blind, randomized, placebo-controlled clinical trials of our group have also shown substantial reduction of the risk of death among critically ill patients with Gram-negative sepsis and ventilator-associated pneumonia with clarithromycin adjunctive treatment [9-11]. We have also recently shown that 28-day mortality of severe CAP was 20.8% among patients treated with a combination of β-lactam and clarithromycin and 33.8% when treated with a combination of β-lactam and azithromycin [12].

Based on the above evidence, it seems likely that treatment of COVID-19 with oral clarithromycin may attenuate the hyper-inflammatory responses of the host and decrease the risk of progression into SRF. We conducted the ACHIEVE trial (<u>A</u>nti-inflammatory <u>C</u>larit<u>h</u>romycin to Improv<u>e</u>SARS-Co<u>V</u> 2 Infection <u>E</u>arly) to investigate whether early administration of oral clarithromycin in patients with upper or lower respiratory tract infection (RTI) due to SARS-CoV-2, may attenuate the inflammatory burden, modulate the immune response of the host and result in early clinical improvement. The primary objective was to investigate the clinical response after early start of treatment. The efficacy of clarithromycin was also compared with a group of propensity-score matched concurrent comparators receiving standard-of-care treatment (SOC) with azithromycin and hydroxychloroquine.

## PATIENTS AND METHODS

### Patient population

ACHIEVE was an open-label non-randomized trial conducted in four study sites in tertiary hospitals in Greece (EudraCT number 2020-001882-36; National Ethics Committee approval 45/20; National Organization for Medicines approval ISO 36/20; ClinicalTrials.gov NCT04398004). Concurrent comparators receiving azithromycin plus hydroxychloroquine/chloroquine were hospitalized at the same time period in five other medical departments of tertiary hospitals of Greece. Written informed consent was provided by the patient or legal representative before screening.

Enrolled patients were adults (age ≥18 years) with confirmed infection by SARS-CoV-2 virus by real-time PCR of nasopharyngeal secretions; and infection of the upper respiratory tract (URTI) or of the lower respiratory tract (LRTI). URTI was defined as the acute presentation of at least two of the following signs in a patient without radiological evidence of lung infection: a) core temperature ≥ 37.5°C; b) new onset of cough; c) chills or rigor; and d) total absolute lymphocyte count less than 1,500/mm^3^. LRTI was defined as the presence of infiltrates compatible with lower respiratory tract infections in chest X-ray or in chest computed tomography accompanied by at least one of the following: a) new onset of cough or worsening cough; b) dyspnea; c) respiratory rales compatible with lung infection; and d) total absolute lymphocyte count less than 1,500/mm^3^. Exclusion criteria were: age below 18 years; intake of any other macrolide and of hydroxychloroquine/chloroquine for the infection under study; ratio of partial oxygen pressure to the fraction of inspired oxygen (pO_2_/FiO_2_) less than 150; need for mechanical ventilation (MV) or non-invasive ventilation under positive pressure (NIV); neutropenia (<1,000/mm^3^); any intake of corticosteroids at a daily dose ≥ 0.4mg/kg prednisone or equivalent the last 15 days; QTc interval at rest electrocardiogram ≥ 500 msec; and pregnancy or lactation.

In order to derive the most appropriate comparators, the inclusion and exclusion criteria of the ACHIEVE trial were applied to all available concurrent comparators.

### Trial interventions

Enrolled patients received one tablet of 500 mg of clarithromycin every 12 hours for 7 days. All other drugs except macrolides and hydroxychloroquine/ chloroquine phosphate were allowed. Study visits were done daily starting from day 1 before start of treatment until day 8 after end of treatment (EOT visit). The test-of-cure (TOC) visit was done on day 14. For patients discharged earlier from hospital, EOT and TOC visits were done by phone calls. Recorded baseline information was severity assessed by APACHE II score, SOFA score and pneumonia severity index (PSI), comorbidities and laboratory information. On each visit follow-up, the state of the patient was assessed and the respiratory symptoms score (RSS) was calculated (see Supplementary methods). Fifteen ml of whole blood was collected on day 1 before start of treatment and on EOT into EDTA-coated tubes and sterile and pyrogen-free tubes for the isolation of peripheral blood mononuclear cells (PBMCs), serum and plasma. PBMCs were isolated after gradient centrifugation over Ficoll (Biochrom, Berlin, Germany) for 20 minutes at 1400g. After three washings in ice-cold PBS pH 7.2, PBMCs were counted in a Neubauer plate with trypan blue exclusion of dead cells. They were then diluted in RPMI 1640 enriched with 2mM of L-glutamine, 500 μg/ml of gentamicin, 100 U/ml of penicillin G, 10 mM of pyruvate, 10% fetal bovine serum (Biochrom) and suspended in wells of a 96-well plate. The final volume per well was 200μl with a density of 2 ×10^6^ cells/ml. PBMCs were exposed in duplicate for 24 hours or 5 days at 37°C in 5% CO_2_ to different stimuli: 10 ng/ml of *Escherichia coli* O55:B5 lipopolysaccharide (LPS, Sigma, St. Louis, USA) or 5×10^5^ colony forming units of heat-killed *Candida albicans*. Following incubation, cells were removed and analysed for flow cytometry. Concentrations of tumour necrosis factor-alpha (TNFα), interleukin (IL)-6, IL-10 and interferon-gamma (IFNγ) were measured in cell supernatants or serum in duplicate by an enzyme immunoassay; the lower limit of detection was 20 pg/ml (Invitrogen, Carlsbad, California, USA). C-reactive protein was measured by nephelometry; the lower limit of detection was 0.5 mg/l.

Nasopharyngeal swabs were collected using the 350C UTM© mini 1ml collection and transport system (COPAN, Italia, S.p.A.) on day 1 before start of treatment, on day 4 and on EOT. The viral load was expressed as the Ct of the reaction positivity and as the relative log2 copies of the *E* gene and of the *RdRp* gene. Viral RNA was extracted from the transport medium using the QIAamp® Viral RNA Mini kit (Qiagen, Hilden, Germany) and all quantified with the Qubit™ RNA HS Assay Kit on a Qubit Fluorometer (Thermo Fisher Scientific Waltham, MA USA). Sequencing libraries were generated using the CleanPlex® SARS-CoV-2 Research and Surveillance Panel powered by SOPHiA (Paragon Genomics, Hayward, CA, USA) using CleanPlex Indexed PCR Primers for Illumina (Paragon Genomics). The generated DNA libraries were loaded in a 300-cycle sequencing cartridge and sequenced on an Illumina MiSeq platform (Illumina, San Diego, CA, USA).

The generated analysis output, in the FASTQ file format, was uploaded to the SOPHiA DDM tool (SOPHiA GENETICS Inc, Boston, MA, USA). A total of 5,598,884 read pairs were generated of which 5,351,919 pairs (95.59%) were successfully mapped. The reference sequence used was MN908947 (NC_045512.2). Four quality indicators generated for all samples (coverage uniformity <90%, effective reads <90%, pool 1 and 2 balance <-1 and >1 log 2 ratio, target covered <95%) allow easy assessment of sequencing results quality. The genome sequences of SARS-CoV-2 were assigned to viral lineages according to the dynamic nomenclature system proposed by Rambaut et al [13] using the SOPHiA DDM tool (https://pangolin.cog-uk.io).

Blood was sampled from concurrent SOC comparators on day 1 before start of treatment and on day 8 for the measurement of biomarkers.

### Outcomes

The primary study endpoint was assessed at EOT and it was a composite endpoint defined differently for patients with URTI and for patients with LRTI. For patients with URTI the primary endpoint was defined as either no need for hospital re-admission in case of earlier discharge or as lack of progression into lower RTI. For patients with LRTI the primary endpoint was defined as any least 50% decrease of the RSS from the baseline provided that the patient has not progressed into SRF or died. SRF was defined as any decrease of the pO_2_/FiO_2_ below 150 mmHg necessitating MV or NIV. This end point was compared between patients who started early and those who started late clarithromycin. The same endpoints were captured for the SOC concurrent comparators. Secondary outcomes were the development of SRF by TOC visit; hospital readmission until TOC; the change of viral load from baseline in respiratory secretions on day 4 and EOT; the change of circulating biomarkers; and the changes of cytokine responses. Study exploratory endpoints were the impact of time delay from the onset of COVID-19 symptoms on the change of the 11-point WHO clinical progression scale [14]; and the association with the lineage of SARS-CoV-2.

Similar definitions of the study endpoints were applied for concurrent SOC comparators. Adverse Events (AE) (Common Terminology Criteria for Adverse Events, version 4.03) and Severe Adverse Events (SAE) (see Supplementary Appendix) were captured.

### Statistical analysis

Qualitative data were presented as percentages with confidence intervals (CI) and quantitative data as means and SD; biomarkers and cytokines were expressed as means and SE. Using the median of the time between symptoms onset and start of clarithromycin, patients were divided into those with an early start and into those with a late start; comparisons were done by the binomial test. Propensity score 1:1 matching was applied to select the best matched comparators to the 90 patients treated with clarithromycin. Matching variables were: age, Charlson’s comorbidity index (CCI), admission PSI; the frequency of URTI and LRTI; and admission C-reactive protein (CRP). Comparisons were performed by the Fisher’s exact test using confirmatory forward stepwise logistic regression analysis (IBM SPSS Statistics v.25.0). Paired comparisons were done by the Wilcoxon’s signed rank test. Any two-sided *P* value <0.05 was statistically significant.

## RESULTS

### Trial conduct

Patients participating in the ACHIEVE open-label trial were enrolled between May 17 2020 and October 8 2020. After propensity matching, 90 concurrent SOC comparators were selected within the same time frame. The propensity score of patients enrolled in the ACHIEVE trial was 0.357 ± 0.094; the respective score of the concurrent SOC comparators was 0.360 ± 0.099 (P: 0.829). The study flow chart is shown in supplementary Figure 1. None of the 180 participants received biological anti-cytokine drugs. The administered dose of azithromycin was 500mg daily for seven days and of hydroxychloquine 500 mg every 12 hours for seven days. Baseline demographics of patients receiving clarithromycin and of concurrent comparators are shown in supplementary Table 1. No difference was found between the two groups.

**Figure 1.**
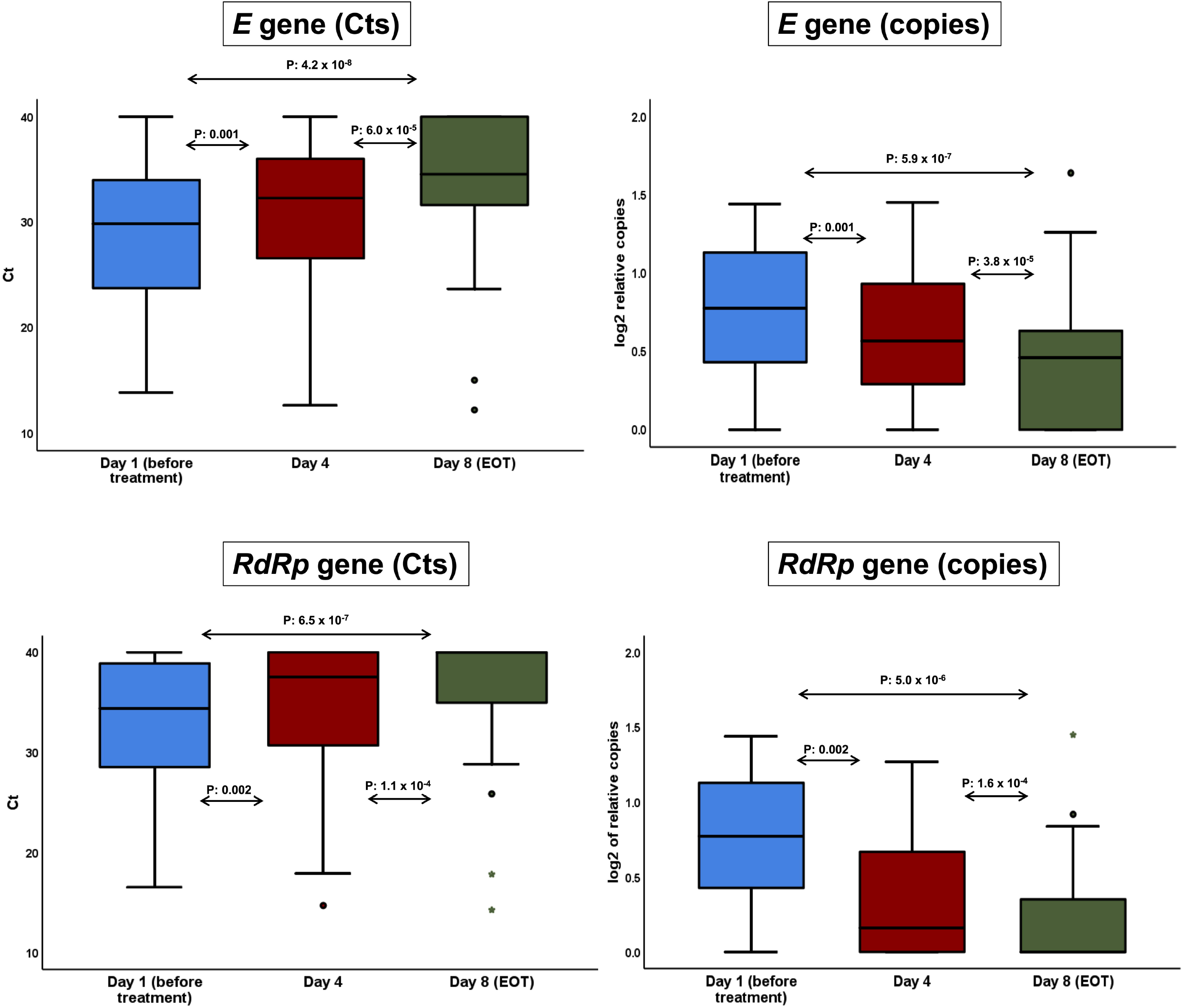
Viral loads of *E* gene and *RdRp* gene in the nasopharynx. Viral load of SARS-CoV-2 was assessed in the nasopharynx of clarithromycin-treated patients over visit using the relative expression of the constitutive *E* gene and of the specific *RdRP* gene. The expression of both genes was expressed as both the absolute time cycle (Ct) of the real-time PCR reaction and the log2 relative copies. The arrows indicate the comparisons; the respective P-value by the Wilcoxon rank-sum test is provided <u>Abbreviation</u> EOT: end-of-treatment

### Study endpoints

The primary endpoint was met in 78 patients treated with clarithromycin (86.7% 95% CIs 78.1-92.2%). Using the median of distribution, patients treated with clarithromycin were divided into those who started treatment within the first 5 days from start of symptoms (n= 48, range 1-5 day) and into those who started treatment 6 days or later from start of symptoms (n= 42, range 6-12 days). No differences were found in baseline characteristics of the enrolled patients divided into those with early and late start of clarithromycin (Table 1). The primary endpoint was achieved in 91.7% (44 out of 48 patients; 95% CIs 80.4-96.7%) and in 81.4% (34 out of 42 patients; 95%CIs 66.7-90.0%) respectively (P: 0.021). This finding was confirmed after multivariate logistic regression analysis showing early start of clarithromycin to be the only independent variable to be positively associated with the favourable primary endpoint (Table 2).

**Table 1.**
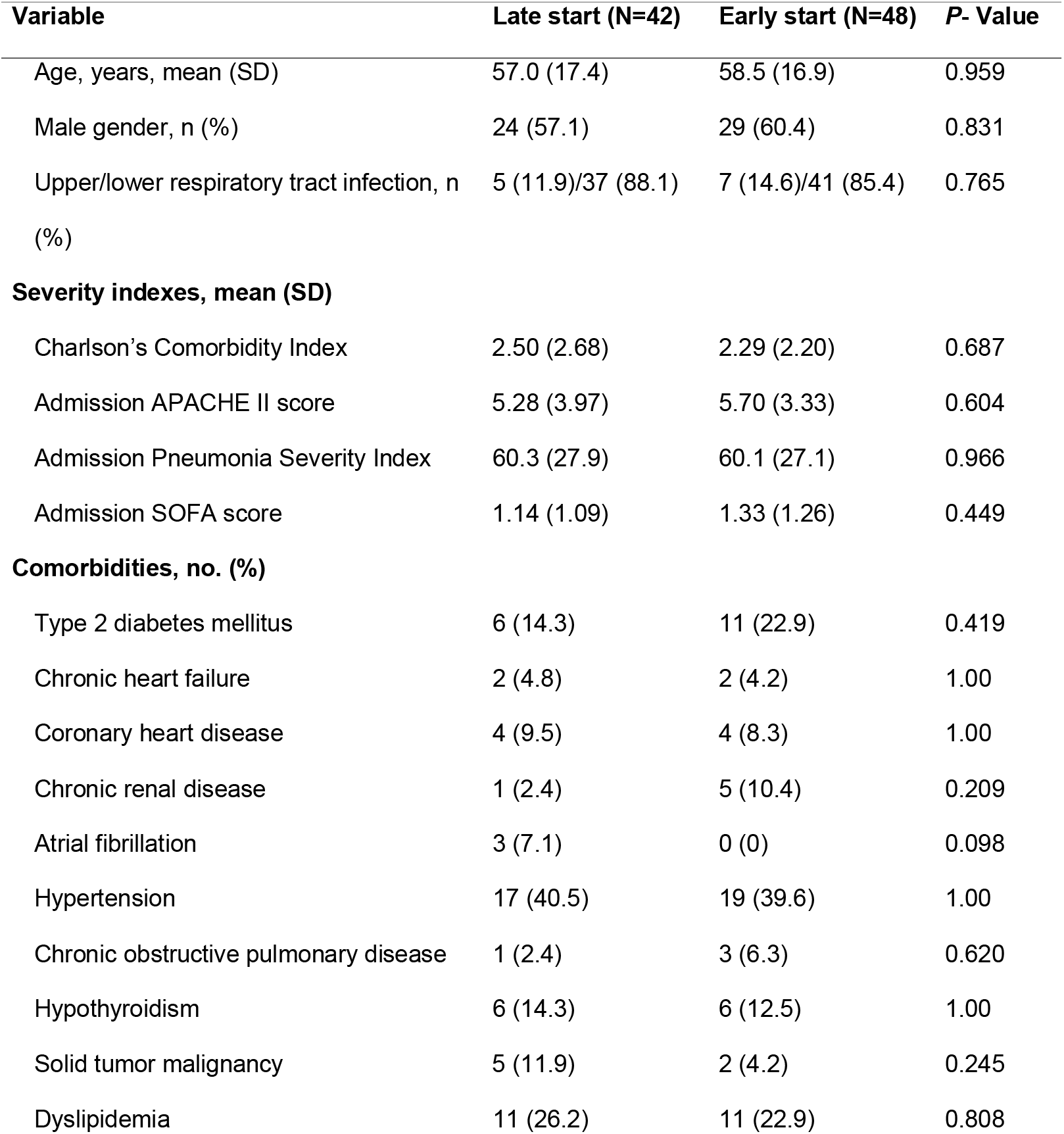
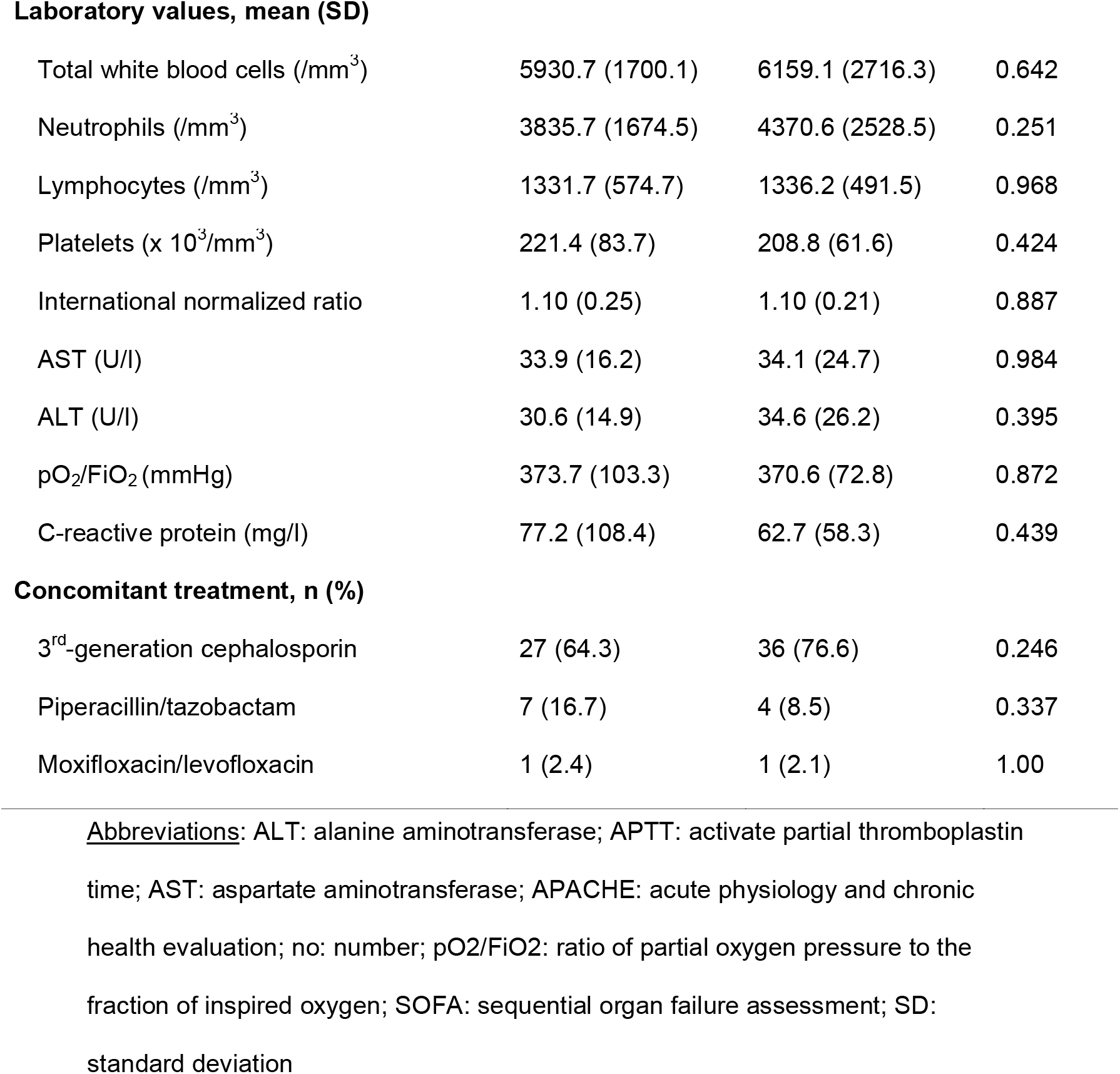
Baseline lab values and treatment modalities for participants in the ACHIEVE trial. Patients are divided onto those who started early treatment with clarithromycin (5 days or less from symptoms onset) and into those who started late treatment with clarithromycin (6 days or more from symptoms onset)

| Variable | Late start (N=42) | Early start (N=48) | P- Value |
| --- | --- | --- | --- |
| Age, years, mean (SD) | 57.0 (17.4) | 58.5 (16.9) | 0.959 |
| Male gender, n (%) | 24 (57.1) | 29 (60.4) | 0.831 |
| Upper/lower respiratory tract infection, n (%) | 5 (11.9)/37 (88.1) | 7 (14.6)/41 (85.4) | 0.765 |
| **Severity indexes, mean (SD)** |  |  |  |
| Charlson's Comorbidity Index | 2.50 (2.68) | 2.29 (2.20) | 0.687 |
| Admission APACHE II score | 5.28 (3.97) | 5.70 (3.33) | 0.604 |
| Admission Pneumonia Severity Index | 60.3 (27.9) | 60.1 (27.1) | 0.966 |
| Admission SOFA score | 1.14 (1.09) | 1.33 (1.26) | 0.449 |
| **Comorbidities, no. (%)** |  |  |  |
| Type 2 diabetes mellitus | 6 (14.3) | 11 (22.9) | 0.419 |
| Chronic heart failure | 2 (4.8) | 2 (4.2) | 1.00 |
| Coronary heart disease | 4 (9.5) | 4 (8.3) | 1.00 |
| Chronic renal disease | 1 (2.4) | 5 (10.4) | 0.209 |
| Atrial fibrillation | 3 (7.1) | 0 (0) | 0.098 |
| Hypertension | 17 (40.5) | 19 (39.6) | 1.00 |
| Chronic obstructive pulmonary disease | 1 (2.4) | 3 (6.3) | 0.620 |
| Hypothyroidism | 6 (14.3) | 6 (12.5) | 1.00 |
| Solid tumor malignancy | 5 (11.9) | 2 (4.2) | 0.245 |
| Dyslipidemia | 11 (26.2) | 11 (22.9) | 0.808 |
| **Laboratory values, mean (SD)** |  |  |  |
| Total white blood cells (/mm<sup>3</sup>) | 5930.7 (1700.1) | 6159.1 (2716.3) | 0.642 |
| Neutrophils (/mm<sup>3</sup>) | 3835.7 (1674.5) | 4370.6 (2528.5) | 0.251 |
| Lymphocytes (/mm<sup>3</sup>) | 1331.7 (574.7) | 1336.2 (491.5) | 0.968 |
| Platelets (x 10<sup>3</sup>/mm<sup>3</sup>) | 221.4 (83.7) | 208.8 (61.6) | 0.424 |
| International normalized ratio | 1.10 (0.25) | 1.10 (0.21) | 0.887 |
| AST (U/l) | 33.9 (16.2) | 34.1 (24.7) | 0.984 |
| ALT (U/l) | 30.6 (14.9) | 34.6 (26.2) | 0.395 |
| pO<sub>2</sub>/FiO<sub>2</sub> (mmHg) | 373.7 (103.3) | 370.6 (72.8) | 0.872 |
| C-reactive protein (mg/l) | 77.2 (108.4) | 62.7 (58.3) | 0.439 |
| **Concomitant treatment, n (%)** |  |  |  |
| 3<sup>rd</sup>-generation cephalosporin | 27 (64.3) | 36 (76.6) | 0.246 |
| Piperacillin/tazobactam | 7 (16.7) | 4 (8.5) | 0.337 |
| Moxifloxacin/levofloxacin | 1 (2.4) | 1 (2.1) | 1.00 |

Abbreviations: ALT: alanine aminotransferase; APTT: activate partial thromboplastin time; AST: aspartate aminotransferase; APACHE: acute physiology and chronic health evaluation; no: number; pO<sub>2</sub>/FiO<sub>2</sub>: ratio of partial oxygen pressure to the fraction of inspired oxygen; SOFA: sequential organ failure assessment; SD: standard deviation

**Table 2.** Univariate and multivariate logistic regression analysis of baseline variables associated with the achievement of the study primary endpoint in the ACHIEVE trial.

| Variable | Incidence of primary endpoint |  | Univariate analysis |  | Multivariate analysis |  |
| --- | --- | --- | --- | --- | --- | --- |
|  | No (total=12) | Yes (total=78) | OR (95% CIs) | P | OR (95% CIs) | P |
| Start of clarithromycin within the first 5 days for symptoms onset, n (%) | 4 (33.3) | 44 (56.4) | 3.49 (0.85-14.34) | 0.082 | 6.62 (1.19-32.63) | 0.030 |
| CCI, mean (SD) | 4.00 (2.95) | 2.14 (2.25) | 0.76 (0.60-0.96) | 0.019 | * |  |
| Admission APACHE II score, mean (SD) | 8.00 (3.13) | 5.12 (3.79) | 0.84 (0.72-0.97) | 0.021 | * |  |
| Admission SOFA score, mean (SD) | 2.16 (1.11) | 1.10 (1.13) | 0.51 (0.31-0.84) | 0.008 | 0.39 (0.22-0.68) | 0.001 |
| Admission PSI, mean (SD) | 80.7 (20.7) | 57.0 (26.9) | 0.97 (0.95-0.99) | 0.008 | * |  |
| Lymphocytes/mm<sup>3</sup>, mean (SD) | 1126.9 (432.3) | 1366.5 (536.9) | 1.00 | 0.148 |  |  |
| CRP, mg/l, mean (SD) | 110.1 (180.2) | 63.7 (61.4) | 0.99 (0.99-1.00) | 0.094 | * |  |

\*variable did not enter the equation after three steps of the multivariate model
Abbreviations APACHE: acute physiology and chronic health evaluation; CCI: Charlson's comorbidity index; CI: confidence interval;
CRP: C-reactive protein; OR: odds ratio; PSI: pneumonia severity index; SOFA: sequential organ failure assessment

RSS was significantly decreased within the first 24 hours from start of clarithromycin (supplementary Figure 2).

**Figure 2.**
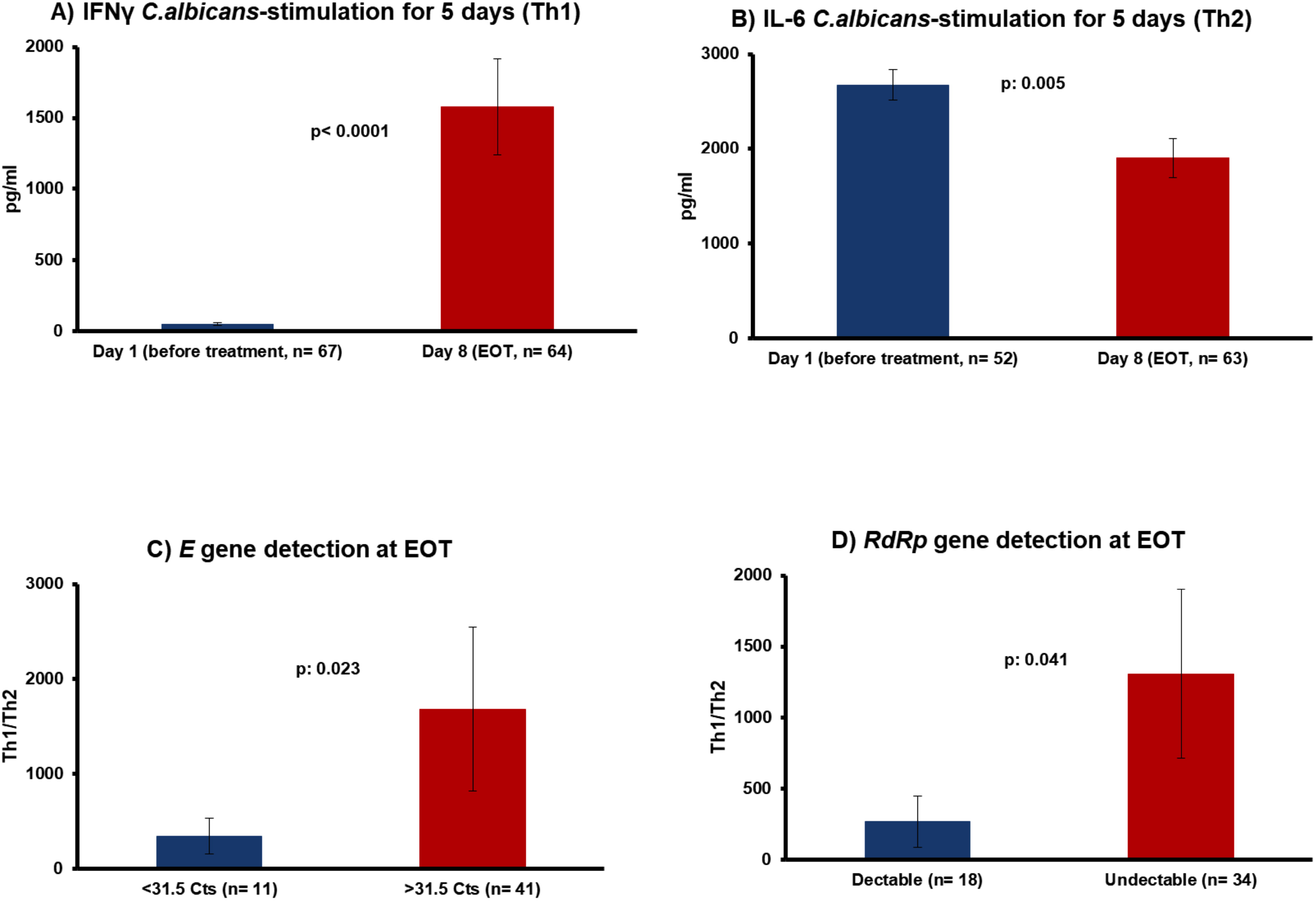
The increase of Th1 response at the end-of-treatment as the proposed mechanism of action of clarithromycin. Production of A) interferon-gamma (IFNγ) reflecting Th1 response; and of B) interleukin (IL)-6 reflecting Th2 response from isolated peripheral blood mononuclear cells following stimulation with heat-killed *Candida albicans* before start of treatment with clarithromycin and at end-of treatment (EOT) with clarithromycin. Comparison of the Th1/Th2 ratio of PBMCs at the EOT between patients with high and low viral load of SARS-CoV-2 on the same treatment visit. The viral load is expressed as the Ct of positivity of the real-time PCR separately for C) the *E* gene and for D) the *RdRp* gene. The N values in parenthesis refer to the existing number of patients analysed. The P-values of comparisons by the Mann-Whitney U-test are provided.

The change of baseline (before start of the study drug) viral load on day 4 of treatment with clarithromycin and at the EOT visit with clarithromycin are shown in Figure 1. The time of Ct to positivity was increased over study visits so as to be considered as indirect index of the decrease of the copies of *E* gene and *RdRp* gene. The relative log2 copies of both genes were decreased over-time.

NGS analysis of viral isolates did not detect any single nucleotide variant (SNV) in either the *E* gene or in the *RdRp* gene. SARS-CoV-2 viral isolates could be determined following quality control amputation in isolates coming from 49 patients. In total, analysis included 32 viral isolates of the B1.1 lineage and 17 isolates of the non-B.1.1 lineage. Patients starting early clarithromycin had an achievement of the positive primary endpoint of treatment response similar for both types of viral lineage. However, among patients starting late clarithromycin the achievement of the positive treatment response was significantly greater among those infected by the non-B1.1 SARS-CoV-2 viral isolates (Table 3).

**Table 3.** Association between delay start of clarithromycin and onset of symptoms and lineage of SARS-CoV-2 viral isolate and achievement of the primary treatment outcome.

| Achievement of primary endpoint | Non-B1.1 lineage*, n (%) | B1.1 lineage, n (%) | P-value | OR (95%CI) |
| --- | --- | --- | --- | --- |
| Early start of clarithromycin ($\leq 5$ days) | | | | |
| No | 1 (14.3) | 2 (9.5) | 1.00 | 1.58 |
| Yes | 6 (85.7) | 19 (90.5) |  | 0.12-20.69 |
| Late start of clarithromycin ($> 5$ days) | | | | |
| No | 0 (0) | 5 (45.5) | 0.035 | NC** |
| Yes | 10 (100) | 6 (54.5) |  |  |

\*Isolates were matched as B1.1 (n= 30, 61.2%); B1.1. with 75% matching to the parent B1.1 (n= 2, 4.1%); B1.1. with 50% matching to the parent B1.1 (n= 8, 16.3%); B1.1.2 (n= 3; 6.1%); B1 (n=2; 4.1%), B1.1.25 (n= 2; 4.1%), B1.1.10 (n=1, 2.1%); and B1.1.70 (n=1, 2.1%). Viral isolates B1.1 and B1.1 with 75% matching to the parent B1.1 were analysed together as viral isolates of the B1.1 lineage.
\*\*NC: cannot be calculated
Abbreviations CI: confidence intervals; OR: odds ratio

Clarithromycin treatment was associated with increased Th1 responses reflected from the increase of the targeted production of IFNγ using stimulation with heat-killed *C*.*albicans* for 5 days (Figure 2A). It was also associated with an attenuation of the Th2 responses as reflected by the targeted production of IL-6 (Figure 2B). The ratio of Th1/Th2 response from PBMCs at the EOT was greater among patients with lower viral load (Figures 2C and 2D). On the contrary, no change of the monocyte function was found (supplementary Figure 3).

**Figure 3.**
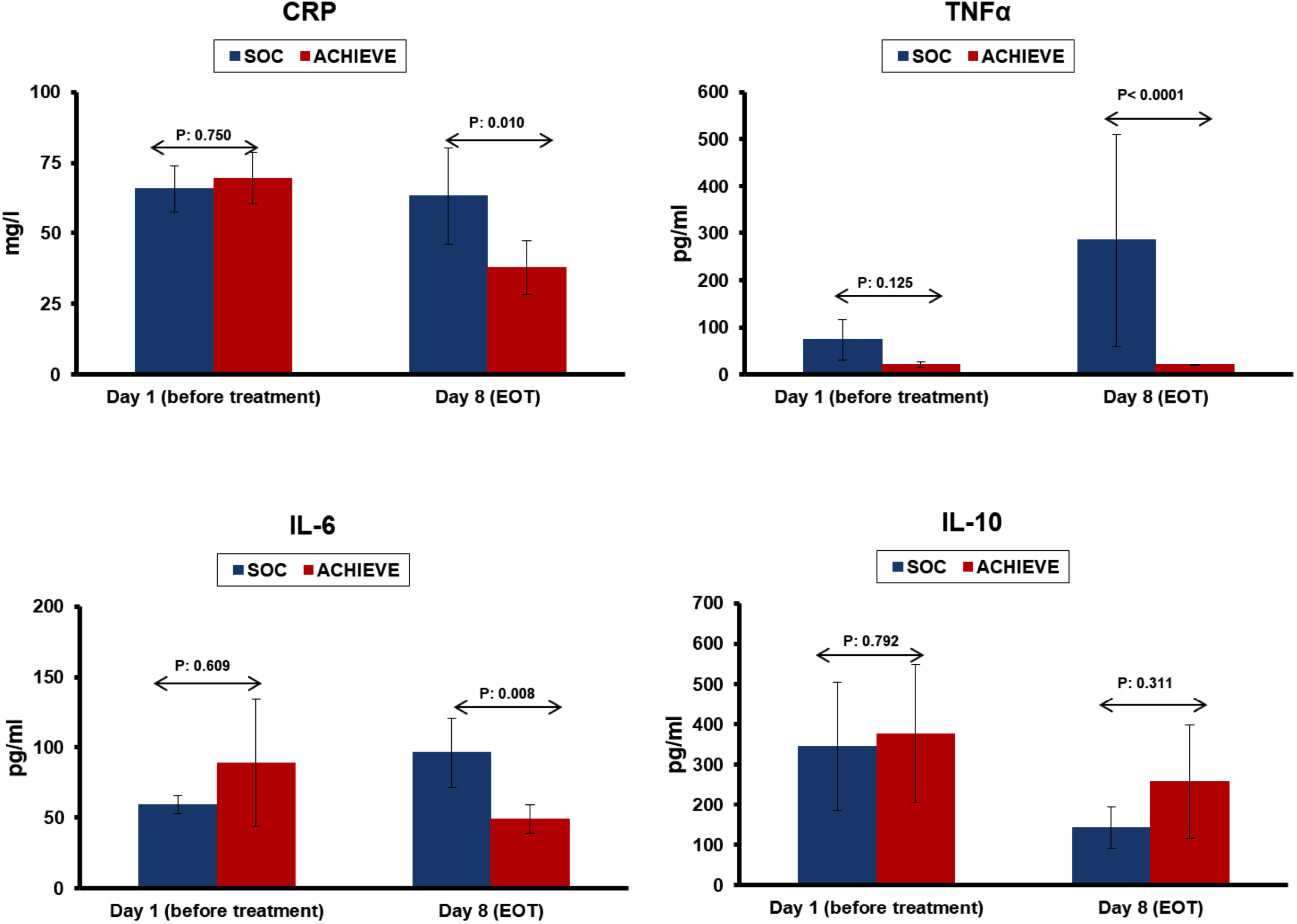
Attenuation of pro-inflammatory responses in COVID-19 following treatment with clarithromycin. Serum concentrations of C-reactive protein (CRP), of tumour necrosis factor-alpha (TNFα) and of interleukin (IL)-6 were measured before start of treatment and at the end-of-treatment visit (EOT) of day 8 among patients treated with clarithromycin (ACHIEVE trial) and among concurrent standard-of-care (SOC) comparators. The P-values of comparisons by the Mann-Whitney U test are provided. <u>Abbreviation</u> EOT: end-of-treatment

The selection of SOC concurrent comparators was appropriate and no differences in baseline characteristics were found (supplementary Table 1). The primary endpoint was met in 66 patients among the concurrent SOC comparators (73.3%; 95%CIs; 63.4-81.4%) and this was significantly lower than patients treated with clarithromycin. When concurrent SOC comparators and patients treated with clarithromycin were analyzed as one cohort, clarithromycin treatment and nine other variables showed an association with the endpoint in univariate logistic regression. However, in a multivariate step-wise logistic regression analysis, only three variables retained a robust independent association with the endpoint (Table 4), including clarithromycin treatment (odds ratio 3.30; 95% CIs 1.10-9.87; P: 0.033).

**Table 4.** Univariate and multivariate logistic regression analysis of baseline variables associated with the achievement of the study primary endpoint. In the analysis patients enrolled in the ACHIEVE trial and concurrent standard-of-care comparators are analysed together.

| Variable | Incidence of primary endpoint |  | Univariate analysis |  | Multivariate analysis |  |
| --- | --- | --- | --- | --- | --- | --- |
|  | No (total=36) | Yes (total=144) | OR (95% CIs) | P- Value | OR (95% CIs) | P- Value |
| Clarithromycin treatment, n (%) | 12 (33.3) | 78 (54.2) | 2.36 (1.09-5.09) | 0.039 | 3.30 (1.10-9.87) | 0.033 |
| CCI, mean (SD) | 3.30 (2.50) | 1.84 (2.07) | 0.77 (0.66-0.89) | 0.001 | * |  |
| Admission APACHE II score, mean (SD) | 7.52 (3.18) | 4.82 (3.67) | 0.82 (0.75-0.92) | <0.0001 | * |  |
| Admission SOFA score, mean (SD) | 2.60 (1.55) | 0.99 (1.03) | 0.39 (0.27-0.54) | <0.0001 | 0.38 (0.25-0.59) | <0.0001 |
| Admission PSI, mean (SD) | 86.8 (27.4) | 58.3 (24.9) | 0.96 (0.95-0.97) | <0.0001 | 0.96 (0.94-0.98) | 0.001 |
| Lymphocytes/mm<sup>3</sup>, mean (SD) | 1052.8 (581.8) | 1299.9 (575.3) | 1.01 (1.00-1.02) | 0.023 | * |  |
| Hypertension, n (%) | 18 (50.0) | 45 (31.3) | 0.45 (0.21-0.95) | 0.037 | * |  |
| Coronary heart disease, n (%) | 7 (19.4) | 7 (4.9) | 0.21 (0.07-0.65) | 0.009 | * |  |
| Atrial fibrillation, n (%) | 6 (16.7) | 1 (0.7) | 0.04 (0.00-0.30) | <0.0001 | * |  |
| CRP, mg/l, mean (SD) | 104.9 (115.8) | 58.2 (68.4) | 0.99 (0.99-0.99) | 0.005 | * |  |

\*variable did not enter the equation after three steps of the multivariate model
Abbreviations APACHE: acute physiology and chronic health evaluation; CCI: Charlson's comorbidity index; CI: confidence interval; CRP: C-reactive protein; OR: odds ratio; PSI: pneumonia severity index; SOFA: sequential organ failure assessment

Clarithromycin treatment was associated with attenuation of pro-inflammatory responses. On day 8 the serum concentrations of C-reactive protein (CRP), tumour necrosis factor-alpha (TNFα) and interleukin (IL)-6 were lower among patients treated with clarithromycin than among concurrent SOC comparators. No differences were found in IL-10 (Figure 3).

At the TOC visit, the incidence of SRF was 12.2% (n= 11; 95%CIs 6.9-20.6%) among patients treated with clarithromycin. This was 26.7% (n= 24; 95%CIs 18.6-36.6%) among SOC concurrent comparators (P: 0.023). Multivariate logistic regression analysis revealed that the only independent variable associated with protection from SRF at TOC was the intake of clarithromycin (odds ratio 0.22; 95% CIs 0.06-0.79; P: 0.022; supplementary Table 2).

The benefit coming from the early start of clarithromycin was reflected on the exploratory outcome of the 11-point WHO CPS by day 28. The decrease of points of the scale of symptoms was significantly greater among early starters of clarithromycin (supplementary Figure 4).

### Safety

The serious adverse events (SAEs) and the non-serious AEs that were captured during the study period of 14 days are listed in supplementary Table 3. Reported events probably or possibly-related to the study drug with incidence greater than 1% were: increase of aminotransferases (Grade I 18.8%; Grade II 1.1%); diarrhea (mild 12.1%; moderate 1.1%); mild vomiting in 1.1%, Grade I increase of bilirubin in 2.22%; and mild allergic reaction in 1.1%). No drug-related SAEs were reported.

## DISCUSSION

Based on previous experience with clarithromycin [9-12], we sought to investigate its potential in reducing the hyper-inflammatory response in moderate COVID-19 and contribute to better clinical outcomes than currently available treatment approaches. Analysis revealed that clarithromycin treatment was associated with a significantly greater clinical benefit at the EOT visit of 86.7%, as compared with 73.3% achieved by the concurrent SOC comparators. This early benefit shown at the end of the 7-day course with clarithromycin was further reflected in the incidence of SRF at the TOC visit of day 14, when treatment with clarithromycin provided 78% relative decrease of the risk for SRF compared to the SOC regimen. The benefit was pronounced for patients starting clarithromycin within the first 5 days from the onset of Covid-19-related symptoms. The benefit of clarithromycin was also linked with decrease of CRP and IL-6 at the EOT.

Emerging data suggest that active virus replication and persistence in the upper respiratory tissue are associated with the severity of the illness [15, 16]. Patients treated with clarithromycin showed remarkable reduction of the viral load in the nasopharynx which was associated with clinical improvement. At the end of treatment with clarithromycin, the Th1 cell responses were increased whereas the Th2 cell responses were attenuated. The Th1 cells are responsible for the containment of the viral replication in viral infections. Specifically, in COVID-19 the inhibition of the Th1-mediated immune responses has a critical impact on the viral replication and the severity of the illness [17]. Patients with lower viral load at the EOT visit had greater Th1/Th2 ratio as this was expressed by modulation of the production of IFNγ and of IL-6 by circulating lymphocytes.

The findings of the ACHIEVE study should be interpreted in the light of certain limitations. The study was designed in mid-March 2020 at the beginning of the pandemic in Greece. It was decided to endorse an open-label and single-arm design in an attempt to include (and benefit) as many patients as possible since no SOC was framed at that time period. The azithromycin plus hydroxychloroquine concurrent comparators were optimally matched without differences in baseline severity and co-administered treatment. When the study was started, most of the patients with COVID-19 hospitalized in the Greek Hospitals were co-administered azithromycin and hydroxychloroquine as an off-label treatment option. In the following months, emerging data showed that hydroxychloroquine does not affect the natural course of COVID-19 [18, 19]. As such, the comparison between the clarithromycin group and the azithromycin plus hydroxychloroquine group should be conceived as comparison between clarithromycin and azithromycin.

In the randomized COALITION trial from Brazil, patients with moderate to severe COVID-19 were randomized to treatment with placebo (n=227), hydroxychloroquine (n=221) and a combination of hydroxychloroquine and azithromycin (n=217). The achievement of the primary endpoint was similar between the three groups of treatment [20]. The same group of investigators performed the COALITION II trial were patients were allocated to treatment with standard-of-care with (n=214) or without azithromycin (n=183). Although the two groups did not differ in the achievement of the primary endpoint after 15 days, patients allocated to azithromycin treatment had a better overall health state after 7 days of treatment [21].

In conclusion, the results of the ACHIEVE trial clearly indicate that clarithromycin treatment in patients with moderate COVID-19 is associated with early clinical improvement and containment of viral load. This is associated with an increase of the ratio of Th1/Th2 response. To the best of our knowledge this is the first study to show the anti-inflammatory impact of clarithromycin on COVID-19. Further studies are needed to better define its future role in mild and moderate COVID-19.

## Data Availability

Date are available from the corresponding author on request.

## Acknowledgements

We would like to thank Hughes associates, Oxford, UK for supporting us with manuscript preparation. The authors would like to thank the patients, families, clinical, laboratory and research staff who contributed to the trial.

## Notes

**Funding statement** This work was supported by Abbott Operation Products AG. Funding body had no role in the design, conduct, analysis and interpretation of data, and decision to publish.

### Competing Interest Statement

P. Panagopoulos has received honoraria from GILEAD Sciences, Janssen, and MSD.
G. Poulakou has received independent educational grants from Pfizer, MSD, Angelini, and Biorad.
H. Milionis reports receiving honoraria, consulting fees and non-financial support from healthcare companies, including Amgen, Angelini, Bayer, Mylan, MSD, Pfizer, and Servier.
E.J. Giamarellos-Bourboulis has received honoraria from Abbott CH, Angelini Italy, InflaRx GmbH, MSD Greece, XBiotech Inc., and BRAHMS GmbH (Thermo Fisher Scientific); independent educational grants from AbbVie Inc, Abbott CH, Astellas Pharma Europe, AxisShield, bioMerieux Inc, Novartis, InflaRx GmbH, and XBiotech Inc; and funding from the FrameWork 7 program HemoSpec (granted to the National and Kapodistrian University of Athens), the Horizon2020 Marie-Curie Project European Sepsis Academy (granted to the National and Kapodistrian University of Athens), and the Horizon 2020 European Grant ImmunoSep (granted to the Hellenic Institute for the Study of Sepsis).
The other authors do not report any conflict of interest.

### Clinical Trial

NCT04398004

### Funding Statement

The study was funded by Abbott Product Operations AG.Funding bodies had no role in the design, conduct, analysis and interpretation of data, and decision to publish.

### Author Declarations

EudraCT number 2020-001882-36; National Ethics Committee approval 45/20; National Organization for Medicines approval ISO 36/20; ClinicalTrials.gov registration NCT04398004. Collection of data from concurrent comparators was approved by the Ethics Committees of AHEPA Thessaloniki General Hospital; ATTIKON University General Hospital; Patras University General Hospital; and Sotiria Athens General Hospital

### Summary of Updates

Data to present the importance of early start of clarithromycin for clinical benefit at the end-of-treatment and for the improvement of the WHO Clinical Progression Scale by day 28

## REFERENCES

1. Giamarellos-Bourboulis EJ, Netea MG, Rovina N, et al. Complex immune dysregulation in COVID-19 patients with severe respiratory failure. Cell Host Microbe 2020; 27: 992–1000.

2. Gautret P, Lagier JC, Parola P, et al. Hydroxychloroquine and azithromycin as a treatment of COVID-19: results of an open-label non-randomized clinical trial. Int J Antimicrob Agents 2020; 56: 105949.

3. Metlay JP, Waterer GW, Long AC, et al. Diagnosis and treatment of adults with community-acquired pneumonia. Am J Resp Crit Care Med 2019; 200: 45–67.

4. GarcÍa Vázquez E, Mensa J, MartÍnez JA, et al. Lower mortality among patients with community-acquired pneumonia treated with a macrolide plus a beta-lactam agent versus a beta-lactam agent alone. Eur J Clin Microbiol Infect Dis, 2005; 24: 190–5.

5. Metersky ML, Ma A, Houck PM, Bratzler DW. Antibiotics for bacteremic pneumonia: improved outcomes with macrolides but not fluoroquinolones. Chest 2007; 131: 466–73.

6. Restrepo MI, Mortensen EM, Waterer GW, Wunderink RG, Coalson JJ, Anzueto A. Impact of macrolide therapy on mortality for patients with severe sepsis due to pneumonia. Eur Resp J 2009; 33: 153–9.

7. Martin-Loeches I, Lisboa T, Rodriguez A, et al. Combination antibiotic therapy with macrolides improves survival in intubated patients with community-acquired pneumonia. Intensive Care Med 2010; 36: 612–20.

8. Nie W, Li B, Xiu Q. β-Lactam/macrolide dual therapy versus β-lactam monotherapy for the treatment of community-acquired pneumonia in adults: a systematic review and meta-analysis. J Antimicrob Chemother 2014; 69: 1441–6.

9. Giamarellos-Bourboulis EJ, Pechère JC, Routsi C, et al. Effect of clarithromycin in patients with sepsis and ventilator-associated pneumonia. Clin Infect Dis 2008; 46: 1157–64.

10. Giamarellos-Bourboulis EJ, Mylona V, Antonopoulou A, et al. Effect of clarithromycin in patients with suspected Gram-negative sepsis: results of a randomized controlled trial. J Antimicrob Chemother 2014; 69: 1111–8.

11. Tsaganos T, Raftogiannis M, Pratikaki M, et al. Clarithromycin leads to long-term survival and cost benefit in ventilator-associated pneumonia and sepsis. Antimicrob Agents Chemother 2016; 60: 3640–6.

12. Kyriazopoulou E, Sinapidis D, Halvatzis S, et al. Survival benefit associated with clarithromycin in severe community-acquired pneumonia: a matched comparator study. Int J Antimiicrob Agents 2020; 55: 105836.

13. Rambaut A, Holmes EC, O’Toole Á, et al. A dynamic nomenclature proposal for SARS-CoV-2 lineages to assist genomic epidemiology. Nat Microbiol 2020; 5: 1403–7.

14. WHO working group on the clinical characterization and management of COVID- 19 infection. A minimal common outcome measure set for COVID-19 clinical research. Lancet Infect Dis 2020; 20: e192–7.

15. Wölfel R, Corman VM, Guggemos W, et al. Virological assessment of hospitalized patients with COVID-2019. Nature 2020; 581: 465–9.

16. Zheng S, Fan J, Yu F, et al. Viral load dynamics and disease severity in patients infected with SARS-CoV-2 in Zhejiang province, China, January-March 2020: retrospective cohort study. BMJ 2020; 369: m1443.

17. Toor SM, Saleh R, Sasidharan Nair V, Taha RZ, Elkord E. T-cell responses and therapies against SARS-CoV-2 infection. Immunology 2021; 162: 30–43.

18. Self WH, Semler MW, Leither LM, et al. Effect of hydroxychloroquine on clinical status at 14 days in hospitalized patients with COVID-19: a randomized clinical trial. JAMA 2020; 324: 2165–76.

19. Kashour Z, Riaz M, Garbati MA, et al. Efficacy of chloroquine or hydroxychloroquine in COVID-19 patients: a systematic review and meta- analysis. J Antimicrob Chemother 2021; 76: 30–42.

20. Cavalcanti AB, Zampieri FG, Rosa RG, et al. Hydroxychloroquine with or without azithromycin in mild-to-moderate Covid-19. N Engl J Med 2020; 38: 2041–52.

21. Furtado RHM, Berwanger O, Fonseca HA, et al. Azithromycin in addition to standard of care versus standard of care alone in the treatment of patients admitted to the hospital with severe COVID-19 in Brazil (COALITION II): a randomized clinical trial. Lancet 2020; 396: 959–67.

