## SUPPLEMENT for "Early start of oral clarithromycin is associated with better outcome in COVID-19 of moderate severity: the ACHIEVE open-label trial"

**Running title: Clarithromycin in COVID-19**

**Konstantinos Tsiakos^1^, Antonios Tsakiris^2^*, Georgios Tsibris^2^*,**

**Pantazis Voutsinas^2^*, Periklis Panagopoulos^3^, Maria Kosmidou^4^,**

**Vasileios Petrakis^3^, Areti Gravvani^1^, Theologia Gkavogianni^2^,**

**Eleftherios Klouras^4^, Konstantina Katrini^2^, Panagiotis Koufargyris^2^, Iro Rapti^4^, Athanassios Karageorgos^2^, Emmanouil Vrentzos^2^, Christina Damoulari^2^,**

**Vagia Zarkada^2^, Chrysanthi Sidiropoulou^5^, Sofia Artemi^2^, Anastasios Ioannidis^6^,**

**Androniki Papapostolou^2^, Evangelos Michelakis^2^, Maria Georgiopoulou^2^, Dimitra-Melia Myrodia^1^, Panteleimon Tsiamalos^5^, Konstantinos Syrigos^1^, George Chrysos^5^, Thomas Nitsotolis^1^, Haralampos Milionis^4^,**

**Garyphallia Poulakou^1^, Evangelos J. Giamarellos-Bourboulis^1^**

*equal contribution

^1^3^rd^ Department of Internal Medicine, National and Kapodistrian University of Athens, Medical School, 115 27 Athens, Greece;

^2^4^th^ Department of Internal Medicine, National and Kapodistrian University of Athens, Medical School, 124 62 Athens, Greece;

^3^2^nd^ Department of Internal Medicine, Democritus University of Thrace, Medical School, 681 00 Alexandroupolis, Greece;

^4^1^st^ Department of Internal Medicine, University of Ioannina, School of Health Sciences, Faculty of Medicine, 455 00 Ioannina, Greece

^5^2^nd^ Department of Internal Medicine, Tzaneion General Hospital of Piraeus, Greece

^6^Department of Nursing, Faculty of Health Sciences, University of Peloponnese, Tripoli, Greece

**METHODS**

| **Symptom** | **Absent (score=0)** | **Mild (score=1)** | **Moderate (score=2)** | **Severe (score=3)** |
| --- | --- | --- | --- | --- |
| **Cough** | No cough or resolution | Cough present but does not interfere with subject’s usual daily activities | Cough present, frequent and does interfere with some of the subject’s usual daily activities | Cough present throughout day and night; limits most of the subjects’ usual daily activities and sleep patterns |
| **Chest pain** | No chest pain or resolution of chest pain | Chest pain present occasionally with deep breathing but does not interfere with subject’s usual daily activities | Chest pain present with normal breaths and does interfere with the subject’s usual daily activities | Chest pain present at rest and/or with shallow breathing; limits most of the subject’s usual daily activities |
| **Shortness of breath/ dyspnea** | No shortness of breath or resolution | Shortness of breath with strenuous activities only but does not interfere with subject’s usual daily activities | Shortness of breath with usual activities and does interfere with the subject’s usual daily activities | Shortness of breath with minimal exertion or at rest; limits most of the subject’s usual daily activities |
| **Sputum** | No coughing up of phlegm/sputum or resolution | Subject coughs up a small amount of phlegm/sputum | Subject coughs up a moderate amount of phlegm/sputum | Subject coughs up a large amount of phlegm/sputum |

### *Score of respiratory symptoms*

### *Adverse events*

Adverse events (AEs) and Serious Adverse Events (SAEs) were collected from baseline until the last patient’s evaluation per protocol. Investigators were monitoring subjects for adverse events and were responsible for recording ALL AEs/SAEs occurring to a patient during the trial.

An adverse event was defined as any undesirable medical occurrence in a subject administered a pharmaceutical product and which did not necessarily have a causal relationship with this treatment. The time relationship was defined from the moment the AE occurs during therapeutic treatment until 5 half-lives after treatment discontinuation. The adverse event may be a sign, a symptom, or an abnormal laboratory finding.

If an adverse event met any of the following criteria, it was considered SAE:

- **Life-threatening situation:** The subject was at risk of death at the time of the adverse event/experience. It does not refer to the hypothetical risk of death.
- **Inpatient hospitalization** or prolongation of existing hospitalization.
- **Persistent or significant disability/incapacity:** This was not intended to include transient interruption of daily activities.
- **Congenital anomaly/birth defects:** Any structural abnormality in subject’s offspring that occurred after intrauterine exposure to treatment.
- **Important medical events/experiences** that may not result in death, be life-threatening, or require hospitalization may be considered a serious adverse event/experience when, based upon appropriate medical judgment, **they may jeopardize the subject and may require medical or surgical intervention to prevent one of the outcomes listed above,** i.e., death, a life-threatening adverse event/experience, inpatient hospitalization or prolongation of existing hospitalization, a persistent or significant disability/incapacity, or a congenital anomaly/birth defect.
- **Spontaneous and elective abortions** experienced by study subject.

**A non-serious adverse event** was any untoward medical occurrence in a patient or subject who is administered a pharmaceutical product, and which does not necessarily have a causal relationship with this treatment. A non-serious adverse event is one that does not meet the definition of a serious adverse event given above.

*Relationship to the drug*

The following definitions were used to assess the relationship of the adverse event to study drug:

- **Probably Related**: The adverse event had a strong time relationship to the drug or relapses if re-induced; another etiology is improbable or clearly less probable.
- **Possibly Related**: The adverse event had a strong time relationship to the drug; alternative aetiology is as probable or less probable.
- **Probably not Related**: The adverse event had a slight or no time relationship to the drug; there is a more probable alternative aetiology
- **Unrelated**: The adverse event was due to an underlying or concomitant disease or to another pharmaceutical product and is not related to the drug (no time relationship and a much more probable alternative aetiology).

**SUPPLEMENTARY FIGURES**

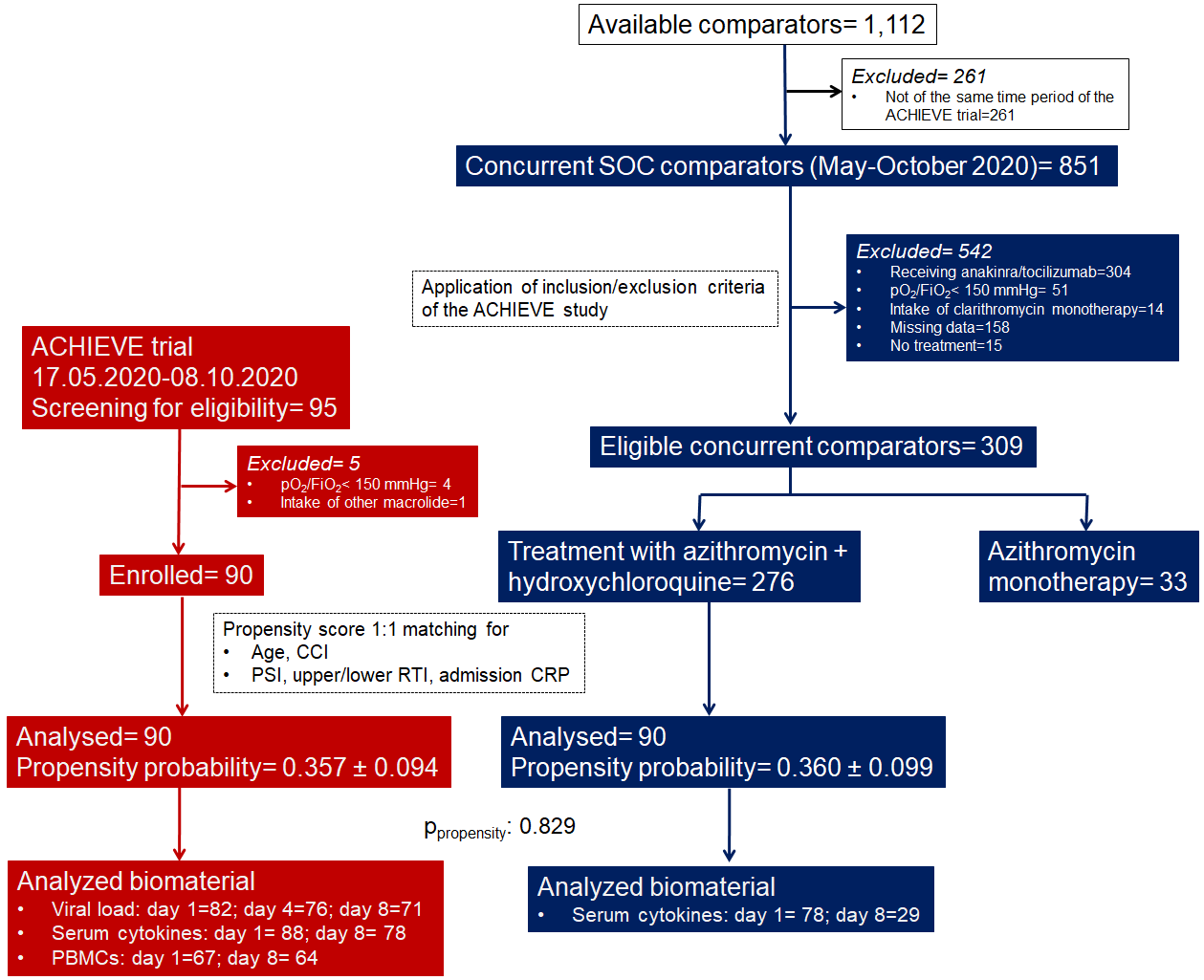

**Supplementary Figure 1 ACHIEVE Trial profile and selection of concurrent standard-of-care (SOC) comparators**

Azithromycin plus hydroxychloroquine/chloroquine comparators were hospitalized at the same time period in five department of Internal Medicine in tertiary hospitals and they were selected in two steps: at the first step after application of the inclusion and exclusion criteria of patients receiving clarithromycin; and at the second step after propensity score matching

Abbreviations CCI: Charlson’s comorbidity index; CRP: C-reactive protein; FiO2: fraction of inspired oxygen; PSI: pneumonia severity index; pO2: partial oxygen pressure; RTI: respiratory tract infection

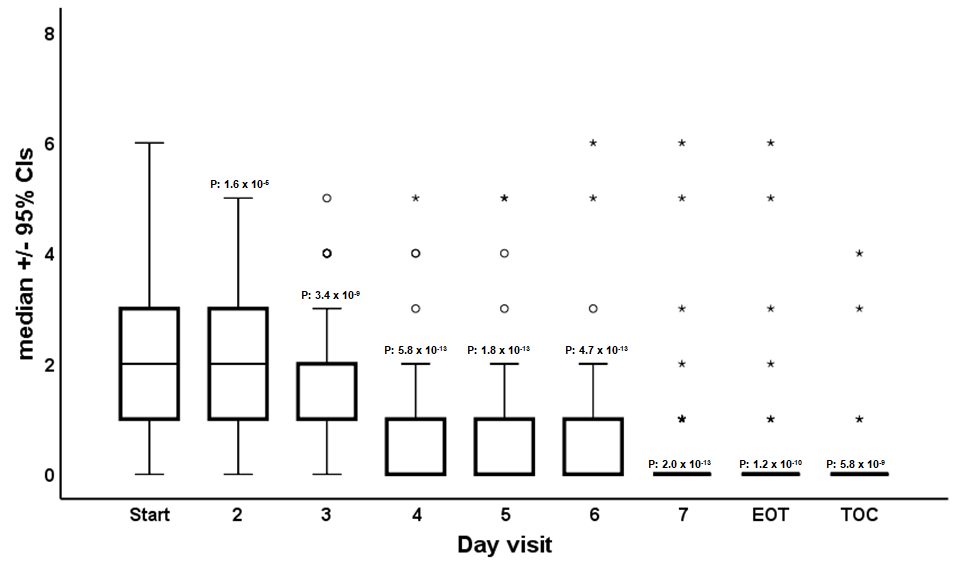

**Supplementary Figure 2 Respiratory symptoms scores on treatment follow-up**

Results are provided only for clarithromycin treated patients. The P-values refer to paired comparisons to day 1 before start of treatment by the Wilcoxon’s test.

Circles denote outliers and asterisk denote extremes. Each time point refers to 78 patients with lower respiratory tract infection by SARS-CoV-2

Abbreviations CI: confidence interval; EOT: End of treatment; TOC: test of cure

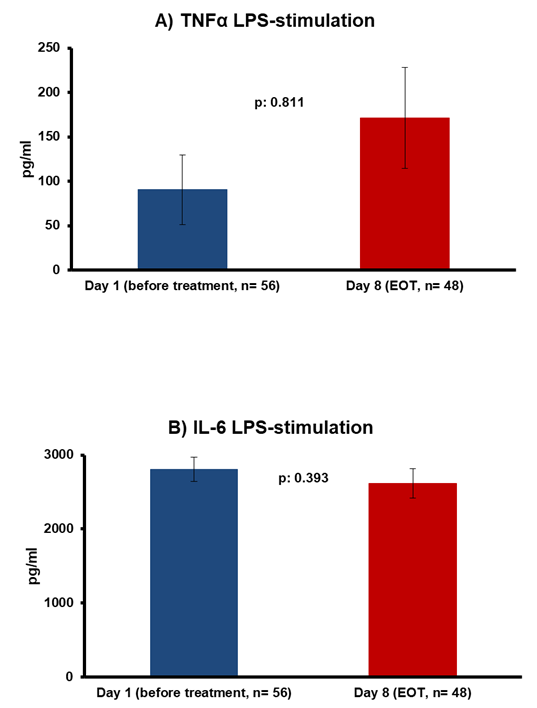

**Supplementary Figure 3 Lack of effect of clarithromycin treatment on monocyte function**

Production of A) tumour necrosis factor-alpha (TNFα); and interleukin (IL)-6 of peripheral blood mononuclear cells (PBMCs) isolated from patients before start of treatment with clarithromycin and one day after end-of-treatment (EOT). PBMCs were stimulated with lipopolysaccharide (LPS) so that produced cytokines reflect monocyte function. The P-values of comparison by the Wilcoxon’s test are provided.

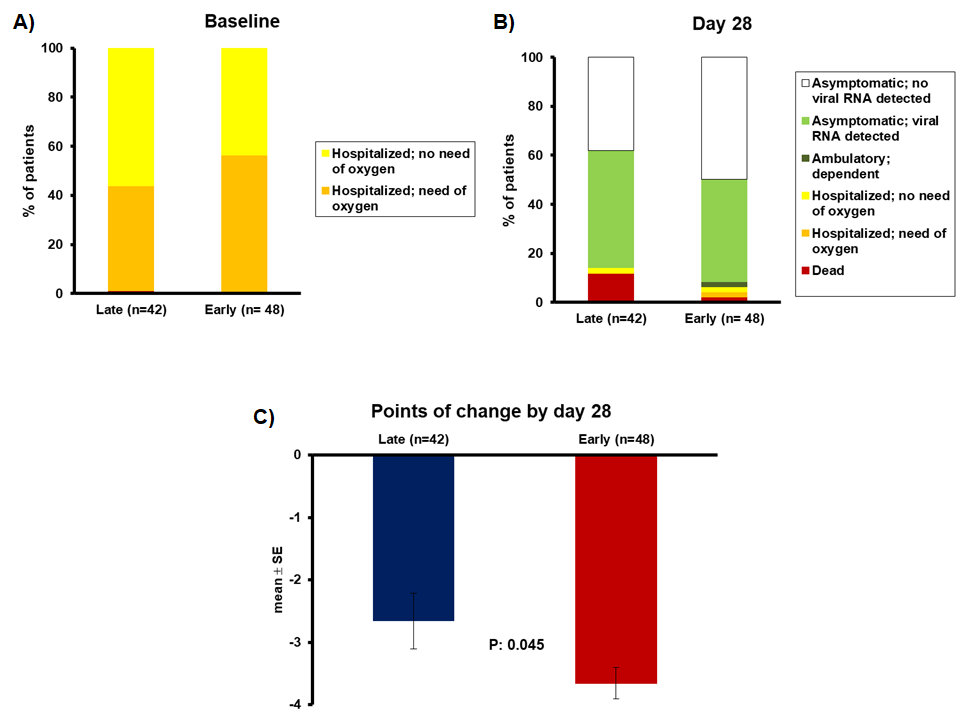

**Supplementary Figure 4 Change of the 11-point WHO clinical progression scale (CPS) after 28 days in association with the time to start of clarithromycin**

Patients are divided into those who started early treatment (the first five days from symptoms onset) and into those who started late treatment (six or more days from symptoms onset). Panel A demonstrates the allocation of patients at the WHO-CPS scale at baseline before start of treatment. Panel B demonstrates the allocation of patients at the WHO-CPS scale after 28 days. Panel C demonstrates the comparative change of points of the WHOC-CPS scale on day 28 from baseline. The p-value of comparison is provided.

**Supplementary Table 1 Baseline lab values and treatment modalities for enrolled patients and parallel standard-of-care (SOC) comparators**

| **Variable** | **SOC comparators (N=90)** | **ACHIEVE trial trial (N=90)** | ***P*- Value** |
| --- | --- | --- | --- |
| Age, years, mean (SD) | 56.7 (16.5) | 56.9 (16.9) | 0.919 |
| Male gender, no. (%) | 57 (63.3) | 53 (58.9) | 0.647 |
| Upper/lower respiratory tract infection, no (%) | 12 (13.3) / 78 (86.7) | 12 (13.3) / 78 (86.7) | 1.00 |
| **Severity indexes, mean (SD)** |  |  |  |
| Charlson’s Comorbidity Index | 1.87 (2.01) | 2.39 (2.42) | 0.125 |
| Admission APACHE II score | 5.31 (3.64) | 5.51 (3.82) | 0.599 |
| Admission Pneumonia Severity Index | 66.0 (28.2) | 62.3 (27.6) | 0.410 |
| Admission SOFA score | 1.37 (1.44) | 1.24 (1.18) | 0.522 |
| **Comorbidities, no. (%)** |  |  |  |
| Type 2 diabetes mellitus | 10 (11.1) | 17 (18.9) | 0.210 |
| Chronic heart failure | 4 (4.4) | 4 (4.4) | 1.00 |
| Coronary heart disease | 6 (6.7) | 8 (8.9) | 0.782 |
| Chronic renal disease | 1 (1.1) | 6 (6.7) | 0.118 |
| Atrial fibrillation | 4 (4.4) | 3 (3.3) | 1.00 |
| Hypertension | 27 (30.0) | 36 (40.0) | 0.211 |
| Chronic obstructive pulmonary disease | 1 (1.1) | 4 (4.4) | 0.368 |
| Hypothyroidism | 11 (12.2) | 12 (13.3) | 1.00 |
| Solid tumor malignancy | 2 (2.2) | 7 (7.8) | 0.169 |
| Dyslipidemia | 16(17.8) | 22 (24.4) | 0.316 |
| **Laboratory values, mean (SD)** |  |  |  |
| Total white blood cells (/mm^3^) | 6114.6 (2869.7) | 6053.9 (2295.2) | 0.876 |
| Neutrophils (/mm^3^) | 4407.7 (2512.9) | 4124.2 (2181.9) | 0.422 |
| Lymphocytes (/mm^3^) | 1167.3 (625.2) | 1334.1 (528.4) | 0.055 |
| Platelets (x 10^3^/mm^3^) | 226.2 (101.9) | 214.7 (72.3) | 0.386 |
| International normalized ratio | 1.12 (0.10) | 1.10 (0.22) | 0.455 |
| aPTT (secs) | 35.83 (28 49) | 34.29 (8.62) | 0.655 |
| AST (U/l) | 38.3 (31.1) | 34.0 (21.1) | 0.280 |
| ALT (U/l) | 34.0 (21.1) | 40.1 (36.5) | 0.105 |
| pO_2_/FiO_2_ (mmHg) | 348.5 (92.1) | 372.0 (87.3) | 0.107 |
| C-reactive protein (mg/l) | 65.6 (78 3) | 69.6 (85.7) | 0.750 |
| **Concomitant treatment, no. (%)** |  |  |  |
| 3^[d^-generation cephalosporin | 61 (67.7) | 63 (70.8) | 1.00 |
| Piperacillin/tazobactam | 13(14.4) | 11 (12.4) | 1.00 |
| Moxifloxacin/levofloxacin | 6 (6.7) | 2 (2.2) | 0.278 |

Abbreviations: ALT: alanine aminotransferase; APTT: activate partial thromboplastin time; AST: aspartate aminotransferase; APACHE: acute physiology and chronic health evaluation; no: number; pO2/FiO2: ratio of partial oxygen pressure to the fraction of inspired oxygen; SOFA: sequential organ failure assessment; SD: standard deviation

**Supplementary Table 2 Univariate and multivariate logistic regression analysis of baseline variables associated with the incidence of severe respiratory failure (SRF) at the test-of-cure visit.** In the analysis patients enrolled in the ACHIEVE trial and concurrent standard-of-care comparators are analysed together.

| **Variable** | **Incidence of SRF** | | **Univariate analysis** | | **Multivariate analysis** | |
| --- | --- | --- | --- | --- | --- | --- |
|  | No (total=145) | Yes (total=35) | OR (95% CIs) | P- Value | OR (95% CIs) | P- Value |
| Clarithromycin treatment, n (%) | 79 (54.5) | 11 (31.4) | 0.38 (0.18-0.84) | 0.017 | 0.22 (0.06-0.79) | 0.022 |
| CCI, mean (SD) | 1.81 (2.02) | 3.45 (2.60) | 1.34 (1.15-1.58) | <0.0001 | * |  |
| Admission APACHE II score, mean (SD) | 4.70 (3.43) | 8.05 (3.74) | 1.27 (1.14-1.41) | <0.0001 | * |  |
| Admission SOFA score, mean (SD) | 0.97 (1.01) | 2.70 (1.53) | 2.83 (1.96-4.09) | <0.0001 | 2.97 (1.78-4.97) | <0.0001 |
| Admission PSI, mean (SD) | 57.5 (23.5) | 90.2 (28.6) | 1.05 (1.03-1.07) | <0.0001 | 1.05 (1.02-1.08) | <0.0001 |
| Lymphocytes/mm^3^, mean (SD) | 1301.4 (574.7) | 1039.7 (579.9) | 0.99 (0.99-1.00) | 0.017 | * |  |
| Coronary heart disease, n (%) | 8 (5.5) | 6 (17.1) | 3.54 (1.14-10.98) | 0.033 | * |  |
| Atrial fibrillation, n (%) | 1 (0.7) | 6 (17.1) | 29.79 (3.45-256.82) | 0.002 | * |  |
| CRP, mg/l, mean (SD) | 57.4 (67.5) | 110.1 (117.1) | 1.00 (1.00-1.01) | 0.003 | 1.00 (1.00-1.01) | 0.024 |

*variable did not enter the equation after four steps of the multivariate model

Abbreviations APACHE: acute physiology and chronic health evaluation; CCI: Charlson’s comorbidity index; CI: confidence interval; CRP: C-reactive protein; OR: odds ratio; PSI: pneumonia severity index; SOFA: sequential organ failure assessment

**Supplementary Table 3 Serious and non-serious adverse events reported by day 14**

| **Event** | **No of patients (%)** | **Relationship with study drug, n (%)** |
| --- | --- | --- |
| **Serious adverse events** | | |
| Mechanical ventilation | 4 (4.4) | None is reported to be related |
| Hypoxemia prolonging hospitalization | 11 (12.2) | None is reported to be related |
| Death | 3 (3.3) | None is reported to be related |
| **Non-serious adverse events** | | |
| Increase of aminotransferases | 56 (62.2) |  |
| Grade I | 52 (57.8) | Probably-related 1 (1.1);  possibly-related 16 (17.7) |
| Grade II | 4 (4.4) | Possibly-related 1 (1.1) |
| Electrolyte abnormalities |  |  |
| Grade I hyperkalemia | 7 (7.7) | None is reported to be related |
| Grade I hypokalemia | 12 (13.3) | None is reported to be related |
| Grade II hypokalemia | 2 (2.2) | None is reported to be related |
| Grade II hyponatremia | 17 (18.9) | None is reported to be related |
| Grade I hyponatremia | 2 (2.2) | None is reported to be related |
| Grade I hypocalcemia | 3 (3.3) | None is reported to be related |
| Gastrointestinal events |  |  |
| Mild diarrhea | 16 (17.1) | Probably-related 3 (3.3);  possibly-related 8 (8.8) |
| Moderate diarrhea | 2 (2.2) | Possibly-related 1 (1.1) |
| Mild vomiting | 2 (2.2) | Possibly-related 2 (2.2) |
| Moderate vomiting | 1 (1.1) | None is reported to be related |
| Grade I bilirubin increase | 3 (3.3) | Possibly-related 2 (2.2) |
| Grade I amylase increase | 7 (7.7) | None is reported to be related |
| Metabolic events |  |  |
| Grade I hyperglycemia | 24 (26.7) | None is reported to be related |
| Grade II hyperglycemia | 7 (7.7) | None is reported to be related |
| Grade I hypoglycemia | 7 (7.7) | None is reported to be related |
| Grade I hypertriglyceridemia | 8 (8.8) | None is reported to be related |
| Blood and lymphatic tissue |  |  |
| Grade I anemia | 16 (17.1) | None is reported to be related |
| Grade II anemia | 2 (2.2) | None is reported to be related |
| Grade I decrease of neutrophils | 2 (2.2) | None is reported to be related |
| Grade II decrease of neutrophils | 1 (1.1) | None is reported to be related |
| Grade I decrease of lymphocytes | 11 (12.2) | None is reported to be related |
| Grade II decrease of lymphocytes | 11 (12.2) | None is reported to be related |
| Grade I decrease of platelets | 6 (6.6) | None is reported to be related |
| Grade II decrease of platelets | 1 (1.1) | None is reported to be related |
| Grade I increase of INR | 3 (3.3) | None is reported to be related |
| Grade II increase of INR | 3 (3.3) | None is reported to be related |
| Grade I increase of aPTT | 1 (1.1) | None is reported to be related |
| Mild allergy | 4 (4.4) | Possibly-related 1 (1.1) |
| Grade I creatinine increase | 8 (8.9) | None is reported to be related |
| Cardiovascular events |  |  |
| Grade I tachycardia | 1 (1.1) | None is reported to be related |
| Grade I bradycardia | 6 (6.6) | None is reported to be related |
| Grade I increase of blood pressure | 1 (1.1) | None is reported to be related |
| Episode of chest pain | 1 (1.1) | None is reported to be related |

Abbreviations aPTT: activated partial thromboplastin time; INR: international normalized ratio
